# Female gender, depressive symptoms, manual job, and higher physical disability predict long term low back pain persistence

**DOI:** 10.1101/2022.02.24.22271451

**Authors:** Luís Antunes Gomes, Ana Maria Rodrigues, Jaime C. Branco, Helena Canhão, Eduardo Brazete Cruz

## Abstract

**Background:** Low Back Pain (LBP) is a long-term health condition with distinct clinical courses. The characterization of these courses together with the identification of prognostic factors of a persistent disabling LBP course has the potential to enable a better identification of patients in high-risk and ultimately allow the development of personalized interventions to change their long-term prognosis. This study aimed to assess the course of chronic LBP (CLBP) over 5 years in a large population-based study, its cumulative impact on disability and health-related quality of life (HRQoL) and the indicators for persistent CLBP course.

**Material and methods:** Active CLBP participants were identified from a representative sample of 10.661 adults randomly recruited from the dwelling population of EpiDoC. Pain, disability and HRQoL were assessed at three time-points. According to their pain symptoms over time, participants were classified as having a persistent or relapsing pain course. A General Linear Model was used to compare mean differences between and within groups. The relation between baseline variables and persistent CLBP was modulated through logistic regression.

**Results:** Among the 1.201 adults with active CLBP at baseline, 634 completed the three time-points of data collection (52.8%) and 400 (63.1%) were classified as having a persistent course. Statistically significant interactions were found between the group and time on disability (*F* (2.126)= 23.78, *p*<0.001) and HRQoL (*F* (2.125)= 82.78, *p*<0.001). In the adjusted model, the persistent course was associated with the disability level (OR: 1.84, CI95% 1.4 to 2.4), presence of depressive symptoms (OR: 1.96, CI95% 1.2 to 3.2), female gender (OR: 1.9, CI95% 1.26-2.87), and having a manual job (OR: 1.46, CI95% 1.02 to 2.1).

**Conclusion:** In the long-term, CLBP patients have distinct clinical courses. Being female, presenting depressive symptoms, having a manual job and a higher disability at baseline predict a CLBP persistent outcome.

## Introduction

Non-specific Low Back Pain (LBP) is one of the most common health problems in societies and is strongly associated with poor self-rated general health [1-4]. It causes severe long-term pain, disability, work absenteeism, and an increased use of healthcare services [1,2].

LBP is traditionally divided into acute (pain of less than 6 weeks), sub-acute (6 to 12 weeks) or chronic (more than 12 weeks) symptom duration [5]. The typical course of acute is initially favorable, i.e., is characterized by a marked reduction in the mean of pain and disability in the first 6 weeks [6]. However, from this point forward, the improvement slows down and the probability of developing a persistent and disabling back pain condition (CLBP – Chronic Low Back Pain) increases for approximately 40% of the patients [6,7]. Some of these patients will recover, but a minority will develop a persistent disabling condition. This minority is responsible for most of the social and health costs associated with LBP [8], to which absenteeism and reduced productivity costs must be added [9].

To better address the individual consequences of persistent disabling LBP on patients and eventually define more appropriate treatment solutions, an improved understanding of its long-term prognosis is needed. Traditionally, LBP episodes have been understood and examined as episodic, recurrent but unrelated [10,11]. Thus, most of the published research is focused on the evaluation of a single episode or presence of pain or disability at specific time-points. It is, therefore, often difficult to know whether a particular study includes individuals with an initial episode of LBP or/and individuals with an exacerbation of a recurrent LBP condition [12,13].

The population-based studies with long-term follow-up (5 to 10 years) published to date have shown that LBP is a persistent long-term condition with a variable clinical course and multiple interrelated episodes [14-21]. Despite differences in the case definition or in the assessment time-points, only 6 to 14% of adults enrolled in these studies reported pain at all specific time-points of the follow-up [18,19]. In contrast, 23 to 35% of the population studied reported that they were “painless at the time of evaluation” [16-18] or had “no day of pain during the previous year” [20]. These results suggest that there is a considerable percentage of individuals reporting pain at a given time who fully recover, while others maintain some level of pain and disability over time, with a small subgroup reporting persistent and potentially disabling pain [16,18-20]. Consequently, the clinical course based on the population mean may not adequately reflect the within-person variation in LBP experienced by individuals, opening the possibility of classifying LBP in clinically relevant subgroups [16,19].

The Epidemiology of Chronic Diseases Cohort Study (EpiDoC), being a population-based study with prolonged follow-up [22-24], produced a set of data that could help identify distinct LBP courses and investigate its impact on disability and quality of life. On the other hand, the presence of distinct clinical courses among LBP patients opens the possibility of identifying prognostic indicators for a persistent LBP course. Although a set of indicators has been associated with the development of persistent and disabling pain [25-30], the majority have emerged from studies that have evaluated pain at a single point of time, with follow-up limited to 1 year [31,32]. The identification of predictors of more disabling LBP course might allow healthcare professionals to better identify the patients at high-risk of persistent disabling symptoms, and eventually change their long-term prognosis through forms of treatment that best fit and respond to their characteristics [16].

The aim of this study was to investigate the presence of different LBP course patterns across 5 years, to examine its cumulative impact on disability and HRQoL and to compare the contribution of socio-demographic, lifestyle, psychosocial and symptom-related indicators to persistent LBP, using data from the EpiDoC.

## Materials and Methods

This study used a longitudinal analysis of active CLBP patients’ data from the EpiDoC. The EpiDoC began with the EpiDoC 1 study (EpiReumaPt) in 2011-13. EpiReumaPt was a population-based study with a representative sample of the Portuguese population, comprised of 10.661 non-institutionalized adults (≥18 years), living in Portugal and able to speak and read the Portuguese language [22,23]. From there, and over a 5-year period, two subsequent time-points of data collection (every 18 months) have been completed, using data based on the same participants [24]. The detailed procedures for the study design and sample selection and recruitment are published elsewhere [23]. Local and regional ethical committees granted ethical approval for this study. All patients gave informed consent after receiving written and oral information about the study [22,23].

This study’s sample consists of all the individuals recruited under the EpiReumaPt study who self-reported active CLBP on the day of the interview [4]. Active CLBP was defined as pain located in the lower back, between the lower thorax and the gluteal folds, that was present most of the time for a minimum period of 90 days, and without a specific cause. Individuals with a diagnosis of other spinal pain (neck or dorsal back pain) or with LBP lasting less than 90 days were excluded [4].

For the purposes of this study, a more stringent case definition was used. Participants who reported “no pain/discomfort” at baseline when answering to question 4 of the Euroqol, 5 dimensions, 3 levels (EQ-5D-3L) *(Regarding pain/discomfort* please indicate which statement best describes your health today?), were excluded. Finally, only those participants who participated in all the time-points of data collection (baseline and time-points 2 and 3) were included in the final analysis.

At baseline, data were collected in face-to-face interviews by trained research assessors (RA) based on a standardized questionnaire which included self-report instruments of disability, health-related quality of life (HRQoL), and anxiety and depressive symptoms. At the follow-up time-points, the RA collected the same self-report measures of disability and HRQoL through telephone interviews. To prevent withdrawals, when a participant was not available, a maximum of 6 additional attempts were made at different times [24].

CLBP course across 5 years was represented by a three-digit variable based on the participants’ answer to question 4 of the EQ-5D-3L: from left to right, the first digit stands for CLBP status at baseline, the second for the pain status at time-point 2, and the third for the pain status at time-point 3.

### Outcome Measures

The primary outcome considered in this study was “persistent LBP” defined as the presence of moderate or extreme pain/discomfort (question 4 of the EQ-5D-3L) at baseline and in all the subsequent time-points of data collection. Additionally, disability status and HRQoL were evaluated through the Portuguese versions of the Health Assessment Questionnaire (HAQ) and the EQ-5D-3L [33,34]. The weight applied to the severity states of the EQ-5D-3L was based on the Portuguese valuation study of the EQ-5D-3L [35].

### Prognostic Indicators

The selection of potential prognostic indicators for the persistent course pattern of CLBP was based on previous published literature [25-30] and covered four domains: socio-demographic (age, gender, employment status, marital status, and education), lifestyle (body mass index, physical exercise, and its frequency), psychosocial (anxiety and depression) and symptom-related indicators (disability and HRQoL).

Anxiety and depression were measured through the Portuguese version of the Hospital Anxiety and Depression Scale (HADS) which is divided into an Anxiety subscale (HADS-A) and a Depression subscale (HADS-D) (the cut-off used for positive anxiety and depression symptoms was >11) [36,37].

### Statistical Analysis

All data analyses were performed using SPSS, Version 24.0 for OX Yosemite (SPSS Inc., Chicago, IL).

Firstly, since complete case analysis may lead to biased results because data are unlikely to be missing completely at random [38], nonresponse bias was assessed by comparing the characteristics of the respondent and non-respondent samples at baseline (time-point 1). A chi-square test and a student’s *t*-test were used to compare the demographic and clinical data of responders and non-responders, as appropriate.

Secondly, and as different CLBP courses were expected, descriptive statistics were planned for different CLBP groups. Baseline characteristics were then compared using the chi-square test and independent *t*-test for categorical and continuous variables, respectively. Differences between groups on disability and HRQoL, over time, were examined using repeated measures analysis of variance (General Linear Model procedure).

Finally, to identify prognostic indicators, associations between the baseline variables and the persistent course of CLBP were modelled through binary logistic regression analysis. A multiple logistic regression was used through backward conditional procedure. All comparisons were adjusted for variables that reveal statistically significant differences in bivariate analysis (*p*<0.20). The non-significant variable with the highest *p*-value was removed in a stepwise manner until all variables had *p*<0.05. The results are presented as crude and adjusted odds ratios (OR) with 95% confidence intervals.

The discriminative ability of the model was assessed through the area under the receiver operating characteristic curve (AUC), ranging from 0.5 (no discrimination) to 1.0 (perfect discrimination) [39]. All *p* values are two-sided, and the significance level was 5%.

## Results

Of the 1.201 selected participants with active CLBP at baseline (time-point 1), 634 (52.8%) completed all the time-points of data collection (complete data on main outcomes - EQ5D index and HAQ). Non-responders at time-point 2 and/or 3 were lost due to unsuccessful contact and for other reasons (participants refused to sign the consent form for follow-up, expressed the wish to leave the study, had an invalid contact or have died) (Fig 1).

**Fig 1.**
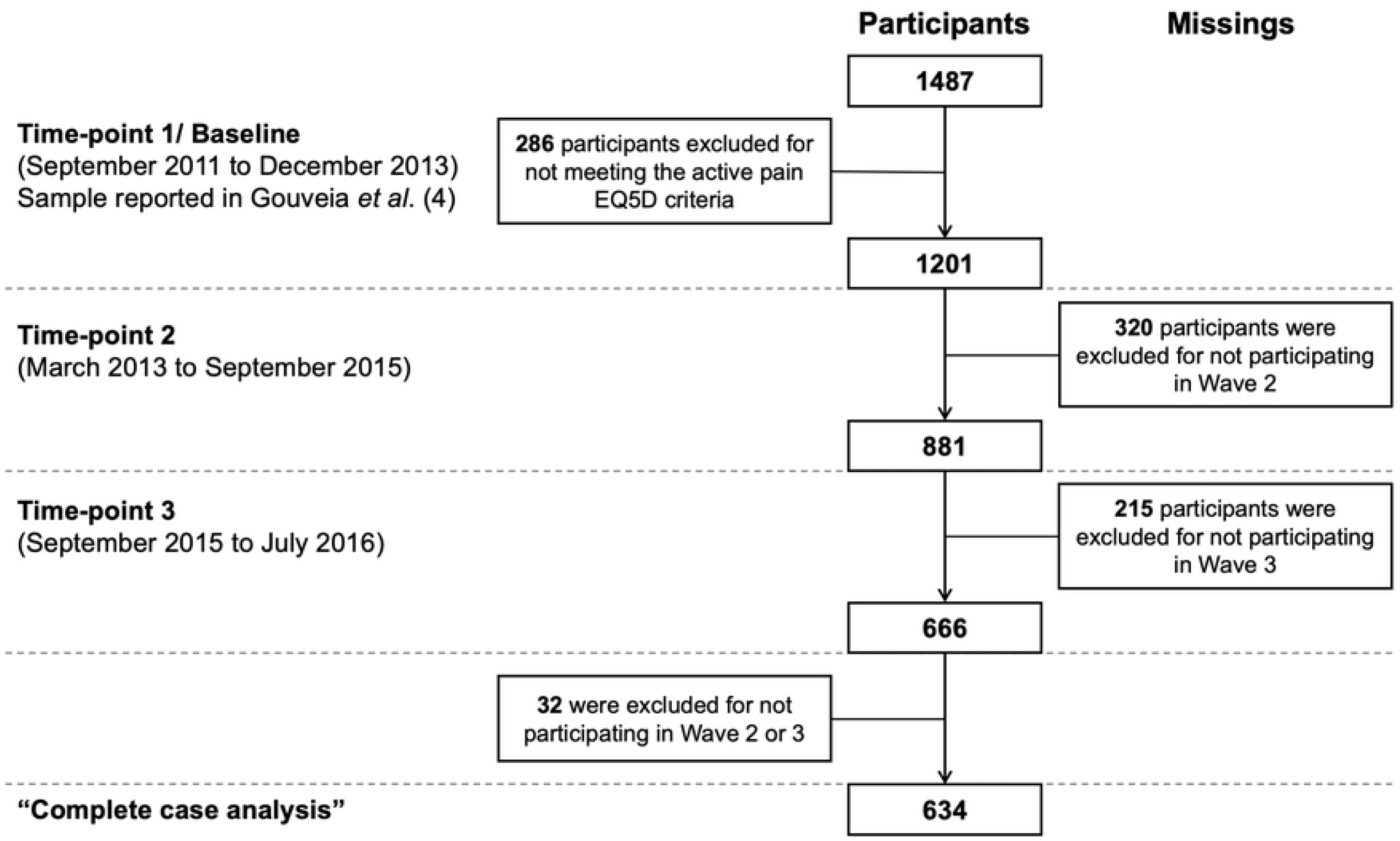
Flow diagram of recruitment.

At baseline, no statistically significant differences in socio-demographic and clinical characteristics were found between responders and non-responders, except for HRQoL (EQ-5D-3L index, *p*=0.005), with responders reporting a statistically significantly lower HRQoL. The participants were aged 61.6 ± 13.6 years, predominantly females (78.4%) and with a low educational level (64.4%). Considering the 3 time-points of data collection, 400 (63.1%) participants were classified as having a persistent LBP course and 234 (36.9%) as having a relapsing LBP course. Of the 234 participants classified with a relapsing LBP course, 63 (26.9%) reported no pain in time-points 2 and 3 (pain/no pain/no pain), 49 (20.9%), an intermittent pattern of pain (pain/no pain/pain), and 122 (52.1%), reported no pain only in the third time-point (pain/pain/no pain). The characteristics of the relapsing and persistent subgroups are presented in Table 1. Statistically significant differences were found between groups for age (*p*=0.008), educational level (*p*=0.001), HRQoL (EQ-5D-3L Index, *p*=0.005) and disability (*p*=0.005). The participants in the persistent subgroup are older and less educated, have higher levels of disability and lower HRQoL (Table 2). Moreover, statistically significant associations were found between gender (*p*≤0.005), anxiety (*p*=0.024) and depressive symptoms (*p*≤0.005), and the type of LBP condition (relapsing or persistent).

**Table 1.**
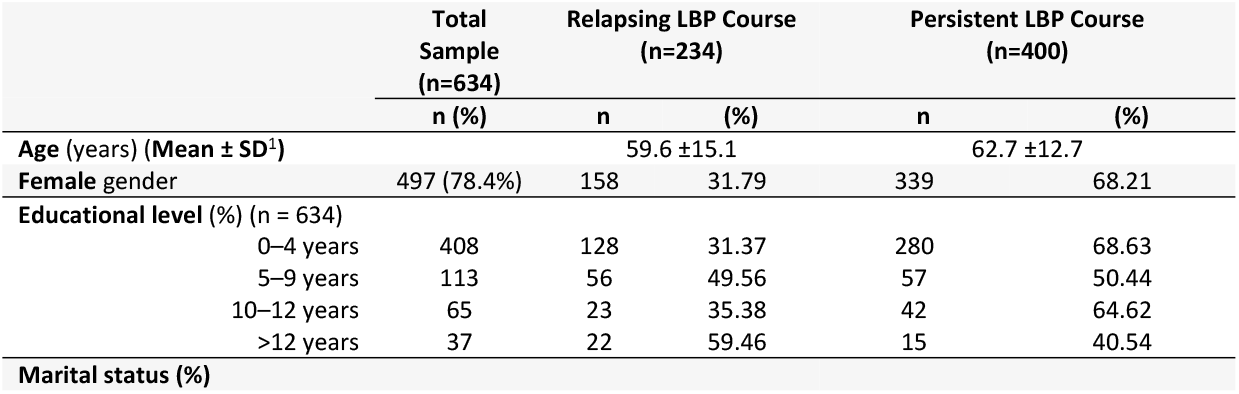
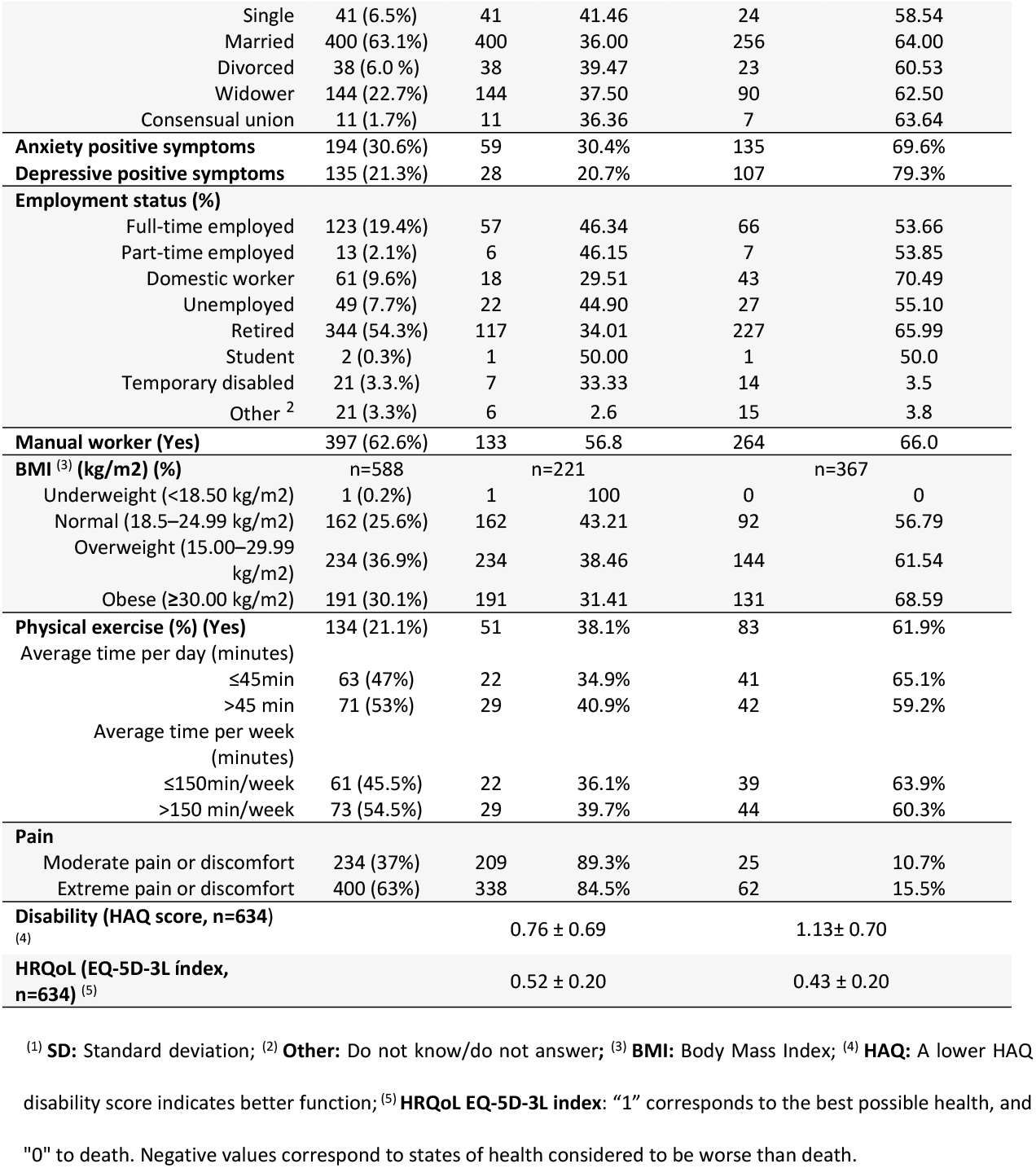
Socio-demographic characteristics of the participants at baseline. Total sample and groups (n=634).

**Table 2.** Health Related Quality of Life and functional status at the different time-points, in “relapsing” and “persistent” LBP subgroups (n=634).

| | | Mean ± SD<br>Relapsing CLBP<br>course | Mean ± SD<br>Persistent CLBP<br>course | p-value |
| --- | --- | --- | --- | --- |
| Time-point<br>1<br>(Baseline) | EQ-5D-3L index<br>(n=634) | 0.52 ± 0.19 | 0.43 ± 0.19 | < .0005 |
| | Score HAQ (n=634) | 0.76 ± 0.69 | 1.13 ± 0.70 | < .0005 |
| Time-point<br>2 | EQ-5D-3L index<br>(n=634) | 0.58 ± 0.28 | 0.30 ± 0.23 | < .0005 |
| | Score HAQ (n=631) | 0.38 ± 0.55 | 0.69 ± 0.80 | < .0005 |
| Time-point<br>3 | EQ-5D-3L index<br>(n=634) | 0.70 ± 0.32 | 0.34 ± 0.24 | < .0005 |
| | Score HAQ (n=631) | 0.55 ± 0.60 | 1.27 ± 0.70 | < .0005 |

<sup>(1)</sup> **HAQ:** A lower HAQ disability score indicates better function; <sup>(2)</sup> **HRQoL EQ-5D-3L index:** "1" corresponds to the best possible health, and "0" to death. Negative values correspond to states of health considered to be worse than death.

### Course of disability and HRQoL between groups, overtime

HRQoL and disability status have improved over time in the “relapsing group”, whereas in the “persistent group” they have remained relatively stable (Table 2). Statistically significant interactions were found between the group (relapsing or persistent) and time (time-point 1 to 3) on disability (*F* (2,1258)=23.779, *p*<0.001), and HRQoL (EQ-5D-3L index), (*F* (2, 1252)=82.779, *p*<0.001). At any time-point there were statistically significant differences on disability (*p*<0.005) and HRQoL (EQ-5D-3L index, *p*<0.005), between groups. Moreover, and over time, an opposite pattern of impact was found between groups; comparing to baseline, HRQoL was statistically significantly higher (*p*<0.001) and disability significantly lower (*p*<0.001) at time-point 3 in the relapsing group, whereas HRQoL statistically significantly decreased (*p*<0.001) and disability significantly increased (*p*<0.001) in the persistent group.

### Prognostic Indicators for the persistent CLBP course

After adjustments, a persistent course of moderate to extreme pain or discomfort was associated with the disability score at baseline, positive depressive symptoms, being female and having a manual job. Participants with higher disability scores at baseline had a higher OR for persistence of pain (1.84, CI95% 1.4-2.4, *p*=0.001). Participants with positive depressive symptoms, manual workers and females have also a higher probability of developing a persistent CLBP course than non-depressive participants (OR: 1.96, CI95% 1.2-3.2, p=0.007), non-manual workers (OR: 1.46, CI95% 1.02-2.1, p=0.04), and males (OR: 1.9, CI95% 1.26-2.87, p=0.002) (Table 3).

**Table 3.** Odds Ratios (95% Confidence Intervals) for the associations between **prognostic indicators and the persistent course of CLBP** (n=400).

| Prognostic indicators | Persistent CLBP Course |  |  |  |  |  |
| --- | --- | --- | --- | --- | --- | --- |
|  | Crude OR | CI 95% | p-value | Adjusted OR | CI 95% | p-value |
| **Socio-demographic Indicators** |  |  |  |  |  |  |
| Age (continuous) | 1.02 | 1.01-1.03 | 0.006** |  |  |  |
| Gender (ref male) | 2.67 | 1.82-3.93 | 0.001** | 1.9 | 1.26-2.87 | 0.002 |
| Educational Level (ref Secondary or higher Education) | 1.45 | 0.94-2.22 | 0.093* |  |  |  |
| Marital Status (ref living alone) | 1.10 | 0.79-1.54 | 0.567 |  |  |  |
| **Psychosocial indicators** |  |  |  |  |  |  |
| Anxiety positive symptoms (Ref no) | 1.51 | 1.05-2.17 | 0.025** |  |  |  |
| Depressive positive symptoms (Ref no) | 2.69 | 1.71-4.23 | 0.001** | 1.96 | 1.21-3.18 | 0.007 |
| Employment status (ref working) | 1.27 | 0.90-1.80 | 0.175* |  |  |  |
| Manual worker (Ref no) | 1.70 | 1.21-2.40 | 0.002** | 1.46 | 1.02-2.1 | 0.040 |
| **Lifestyle Indicators** |  |  |  |  |  |  |
| BMI (Ref Underweight/normal weight) | 1.44 | 0.99-2.08 | 0.055* |  |  |  |
| Physical exercise (Ref yes) | 1.06 | 0.72- 1.58 | 0.756 |  |  |  |
| **Symptom-related Indicators** |  |  |  |  |  |  |
| Pain (Question 4- EQ-5D-3L)<br>(Ref Moderate pain) | 1.53 | 0.93-2.52 | 0.091* |  |  |  |
| Disability (HAQ<sup>4</sup>, 0-24) | 2.19 | 1.71-2.90 | 0.001** | 1.84 | 1.4-2.4 | 0.001 |
| HRQoL (EQ5D<sup>5</sup> Index, 0-1) | 0.74 | 0.03-0.19 | 0.001** |  |  |  |
| **Area Under the Curve (AUC)** |  |  |  | 0.70 | 0.68-0.74 | 0.001 |

\* $P<0.2$ ; \*\* $P<0.05$ ; \*\*\* $P<0.01$ .
<sup>1</sup> **OR**: Odds Ratio; <sup>2</sup> **CI**: Confidence Intervals; **BMI**: Body Mass Index; <sup>(4)</sup> **HAQ**: A lower HAQ disability score indicates better function; <sup>(5)</sup> **HRQoL EQ-5D-3L index**: "1" corresponds to the best possible health, and "0" to death. Negative values correspond to states of health considered to be worse than death.

## Discussion

The aim of this study was to investigate the presence of different LBP symptoms’ courses during a follow-up period of 5 years, to examine its cumulative impact on disability and HRQoL and to quantify the contribution of different prognostic indicators in predicting the long-term persistence of symptoms. To our knowledge, this is the first large prospective study that addresses the long-term course and prognosis of a restricted sample of active CLBP patients. Its results provide evidence for different symptoms’ courses among people classified as chronic patients according to the traditional temporal categorization of LBP, suggesting that classifying LBP simply as acute or chronic is insufficient [5-17], and supporting the current understanding of LBP as a relapsing condition of interrelated episodes of pain [11,14,18-20,40].

In this study, 26.9% of participants reported at least one time-period of complete remission of LBP symptoms. This result is consistent with other long-term population studies’ findings that reported values of pain-free people in all follow-up points between 23 and 35% [18-20]. Contrary to previous studies, the proportion of participants that had reported a persistent symptoms’ course at all follow-up points (63.1%) was substantially higher, with values ranging from 6 to 29% [18-20]. These differences might be explained in part by differences in sample characteristics [7], or the long-standing LBP definition used and data collection methods [18-20,41]. In this study, and since no questions were made to participants regarding their LBP symptoms between time-points, it is possible that the persistent group includes people with continuous mild pain with low impact and people with a fluctuating symptoms course that varies between severe pain episodes with a large impact on their live and pain-free periods [15].

The results obtained regarding the course of disability and HRQoL seem to support this relapsing course of LBP symptoms. Despite statistically significant differences between groups at all follow-up time-points, both groups share a similar relapsing pain pattern over time. Nevertheless, it is important to highlight that statistically significant differences found between the baseline and time-point 3 indicate that disability decreases over time in the relapsing group and increases over time in the persistent group. Comparisons of this study’s findings with other studies are difficult since those designed to describe the long-term course of disability and/or HRQoL are rare.

The present study explored potential prognostic indicators for persistent LBP. Although there is a vast amount of studies dedicated to risk factors for the onset of LBP or concerned with the transition of acute to CLBP, few studies investigated prognostic indicators for persistent CLBP over periods of more than 1 year. In this study, being female, have a manual job, presenting positive depressive symptoms, and reporting disability, increases the chances of a persistent long-lasting LBP. These indicators are consistent with findings reported in other studies [42-46].

Overall, the four indicators identified show a moderate ability to distinguish between relapsing and persistent courses (AUC 0.70). This result limits the capacity of these indicators to identify at presentation patients who are at risk for persistent disabling symptoms, and suggests that other potential prognostic factors (e.g., physical, or psychosocial) should be considered to predict persistent LBP.

### Strengths and Limitations

Strengths of the current study include its long-term prospective design, the large sample size, data on the long-term impact of persistent versus relapsing CLBP courses on disability and HRQoL and the identification of prognostic indicators for the persistent subgroup.

Nevertheless, some methodological limitations must be considered. First, only 52.8% of the baseline sample responded to both follow-ups. Although participants included in this study were similar to those lost, except for HRQoL, the possibility of selection bias and residual confounding cannot be ruled out.

Another limitation already mentioned was the lack of information regarding LBP symptoms between data collection periods. Future studies should seek to enroll patients with an identical starting point of LBP symptoms’ and follow them more frequently over time, to allow a better understanding on whether a certain moment of pain is due to a recurrence episode of pain or a persistent pain course. This distinction will make it possible to better distinguish different courses of pain among CLBP patients and potentiate the design of specific interventions whether to reduce the number of recurrences, in patients with relapsing course, or to promote self-management in patients with a persistent course. Furthermore, and although positive depressive symptoms were significantly associated with persistent LBP, not all potentially important psychological constructs were included in the present study. For example, fear of pain, catastrophizing [47], and illness perceptions were found to be the strongest predictors of poor prognosis in previous research [48].

## Conclusions

The results of this study provide evidence on the long-term course of back pain and suggested that people with CLBP might have different clinical courses ahead and different prognosis in the long-term. It also provides relevant information about how persistent CLBP might affect disability and HRQoL over time. Depressive and female patients that have a manual job and report disability, have higher chances to develop a persistent long-lasting LBP.

## Data Availability

All relevant data are within the manuscript and its Supporting Information files.

## Acknowledgments

The authors team wish to acknowledge the contribution of Professor Carla Nunes for the conceptualization and formal analysis of the current study.

## References

1. Global Burden of Disease, Injury Incidence, Prevalence Collaborators. Global, regional, and national incidence, prevalence, and years lived with disability for 310 diseases and injuries, 1990–2015: a systematic analysis for the Global Burden of Disease Study 2015. Lancet 2016;388:1545–602.

2. Vos T, Flaxman AD, Naghavi M, et al. Years lived with disability (YLDs) for 1160 sequelae of 289 diseases and injuries 1990–2010: a systematic analysis for the Global Burden of Disease Study 2010. Lancet. 2012;380(9859):2163–96.

3. Branco J, Rodrigues AM, Gouveia N, et al. Prevalence of rheumatic and musculoskeletal diseases and their impact on health related quality of life, physical function and mental health in Portugal: results from EpiReumaPt – a national health survey. RMD Open 2016;2:e000166. doi:10.1136/rmdopen-2015-000166.

4. Gouveia N, Rodrigues A, Eusébio M, et al. Prevalence and social burden of active chronic low back pain in the adult Portuguese population: results from a national survey. Rheumatology International 2016;36(2):183–97.

5. van Tulder M, Becker A, Bekkering T, et al. European guidelines for the management of acute nonspecific low back pain in primary care. COST Action B13 2001.

6. Costa LC, Maher CG, Hancock MJ,et al The prognosis of acute and persistent low-back pain: a meta-analysis. CMAJ 2012;184(11):E613–24.

7. Costa LC, Maher CG, McAuley JH, et al. Prognosis for patients with chronic low back pain: inception cohort study. BMJ 2009;339:b3829.

8. Global Burden of Disease 2015 DALYs and HALE Collaborators. Global, regional, and national disability-adjusted life years (DALYs) for 306 diseases and injuries and healthy life expectancy (HALE) for 188 countries, 1990-2013: quantifying the epidemiological transition. Lancet 2015;386:2145–91.

9. Maetzel A, Li L. The economic burden of low back pain: a review of studies published between 1996 and 2001. Best Practice & Research Clinical Rheumatology 2002;16(1):23–30. doi:10.1053/berh.2001.0204.

10. Hartvigsen J, Hancock MJ, Kongsted A, et al. What low back pain is and why we need to pay attention. Lancet 2018;391(10137):2356–67.

11. Dunn, KM, Hesbaek L, Cassidy JD. Low back pain across the life course. Best Practice Research and Clinical Rheumatology, 2013;27(5):591–600. doi: 10.1016/j.berh.2013.09.007.

12. Hancock MJ, Maher CG, Laslett M, et al. Discussion paper: what happened to the “bio” in the bio-psycho-social model of low back pain? European Spine Journal 2011;20(12):2105–10. 3

13. Koes BW, van Tulder MW, Thomas S. Diagnosis and treatment of low back pain. BMJ 2006;332:1430–1434.

14. Hoy D, Brooks P, Blyth F, et al. The epidemiology of low back pain. Best Pract Res Clin Rheumatol 2010;24:769–781.

15. Dunn KM, Campbell P, Jordan KP. Long-term trajectories of back pain: cohort study with 7-year follow-up. BMJ Open 2013;3:e003838. doi:10.1136/bmjopen-2013-003838.

16. Tamcan O, Mannion AF, Eisenring C, et al. The course of chronic and recurrent low back pain in the general population. Pain 2010;150:451–7.

17. Hestbaek L, Leboeuf-Yde C, Manniche C. Low back pain: what is the long-term course? A review of studies of general patient populations. European Spine Journal 2003;12(2):149–65.

18. Kolb E, Canjuga M, Bauer GF, et al. Course of back pain across 5 years: a retrospective cohort study in the general population of Switzerland. Spine 2011;36 (4):E268–73.

19. van Oostrom SH, Verschuren WM, de Vet HC, et al. Ten year course of low back pain in an adult population-based cohort – the Doetinchem Cohort Study. European Journal of Pain 2011;15(9):993–8.

20. Hestbaek L, Leboeuf-Yde C, Engberg M, et al. The course of low back pain in a general population. Results from a 5-year prospective study. Journal of Manipulative Physiological Therapy 2003;26(4):213–9.

21. Itz CJ, Geurts JW, van Kleef M et al. Clinical course of non-specific low back pain: a systematic review of prospective cohort studies set in primary care. European Journal of Pain 2013;17(1):5–15. doi:10.1002/j.1532-2149.2012.00170.x

22. Ramiro S, Canhao H, Branco JC. EpiReumaPt Protocol— Portuguese epidemiologic study of the rheumatic diseases. Acta Reumatológica Portuguesa 2010;35:384–90.

23. Rodrigues AM, Gouveia N, da Costa LP, et al. EpiReumaPt - the study of rheumatic and musculoskeletal diseases in Portugal: a detailed view of the methodology. Acta Reumatológica Portuguesa 2015;40:110–24.

24. Dias S, Rodrigues AM, Gregório MJ, et al. Cohort Profile: The Epidemiology of Chronic Diseases Cohort (EpiDoC). International Journal of Epidemiology 2018;1741–1742j doi: 10.1093/ije/dyy185.

25. Chou R, Shekelle P. Will this patient develop persistent disabling low back pain? JAMA 2010;303:1295–302.

26. Hendrick P, Milosavljevic S, Hale L, et al. The relationship between physical activity and low back pain outcomes: a systematic review of observational studies. European Spine Journal 2011;20:464–74.

27. Pinheiro MB, Ferreira ML, Refshauge K, et al. Symptoms of depression as a prognostic factor for low back pain: a systematic review. Spine Journal 2016;16:105–16.

28. Wertli MM, Eugster R, Held U, et al. Catastrophizing – a prognostic factor for outcome in patients with low back pain: a systematic review. Spine Journal 2014;14:2639–57.

29. Wertli MM, Rasmussen-Barr E, Weiser S, et al. The role of fear avoidance beliefs as a prognostic factor for outcome in patients with nonspecific low back pain: a systematic review. Spine Journal 2014;14:816–36.

30. Wilkens P, Scheel IB, Grundnes O, et al. Prognostic factors of prolonged disability in patients with chronic low back pain and lumbar degeneration in primary care: a cohort study. Spine Journal 2013;38:65–74.

31. Deyo RA, Bryan M, Comstock BA, et al. Trajectories of Symptoms and Function in Older Adults with Low Back Disorders. Spine Journal 2015;40(17):1352–362.

32. Chen Y, Campbell P, Strauss V, et al. Trajectories and predictors of the long-term course of low back pain: cohort study with 5-year follow-up. Pain 2018;159(2):252–260.

33. Fries JF, Spitz P, Kraines RG, et al. Measurement of patient outcome in arthritis. Arthritis & Rheumatology 1980;23:137–145.

34. Ferreira LN, Ferreira PL, Pereira LN. Contributos para a Validação da Versão Portuguesa do EQ-5D. Acta Médica Portuguesa 2013;26(6):664–675.

35. Ferreira LN, Ferreira PL, Pereira LN, et al. The valuation of the EQ-5D in Portugal. Quality of Life Research 2014;23(2):413–423.

36. Zigmond AS, Snaith RP. Hospital anxiety and depression scale (HAD). Acta Psychiatrica Scandinavica 1983;67:361–70.

37. Pais-Ribeiro J, Silva I, Ferreira T, et al. Validation Study of a Portuguese version of the hospital anxiety and depression scale. Psychology, Health & Medicine 2007;12(2):225–237.

38. Sterne JA, White IR, Carlin JB, et al. Multiple imputation for missing data in epidemiological and clinical research: potential and pitfalls. BMJ 2009;338:b2393.

39. Hosmer DW, Lemeshow S. Applied logistic regression. 2nd Edition, New York, John Wiley & Sons, Inc 2000; doi 10.1002/0471722146.

40. Dunn KM, Jordan K, Croft PR. Characterizing the course of low back pain: a latent class analysis. American Journal of Epidemiology 2006;163:754–61.

41. Waxman R, Tennant A, Helliwell P. A prospective follow-up study of low back pain in the community. Spine Journal 2000;25:2085–90.

42. Thomas E, Silman AJ, Croft, PR, et al. Predicting who develops chronic low back pain in primary care: a prospective study. BMJ 1999;318:1662–7.

43. Grotle M, Foster NE, Dunn KM, et al. Are prognostic indicators for poor outcome different for acute and chronic low back pain consulters in primary care? Pain 2010;151:790–797.

44. Carey TS, Garrett JM, Jackman AM. Beyond the good prognosis: examination of an inception cohort of patients with chronic low back pain. Spine Journal 2000;25:115–20.

45. Costa LC, Maher CG, McAuley JH, et al. Prognosis for patients with chronic low back pain: inception cohort study. BMJ 2009;339:b3829.

46. Schiottz-Christensen B, Nielsen GL, Hansen VK, et al. Long-term prognosis of acute low back pain in patients seen in general practice: a 1-year prospective follow-up study. Family Practice 1999;16:223–32.

47. Dunn, KM, Jordan KP, Croft PR. Contributions of prognostic factors for poor outcome in primary care low back pain patients. Eur J Pain 2011;15(3):313–9.

48. Foster NE, Thomas E, Bishop A, et al. Distinctiveness of psychological obstacles to recovery in low back pain patients in primary care. Pain 2010;148:398–406.

